# Alterations in gamma frequency oscillations correlate with cortical tau deposition in Alzheimer’s disease

**DOI:** 10.1101/2023.12.21.23300275

**Authors:** Umberto Nencha, Isotta Rigoni, Federica Ribaldi, Daniele Altomare, Margitta Seeck, Valentina Garibotto, Serge Vulliémoz, Giovanni B. Frisoni

**Author notes:** <u>Corresponding author:</u> Dr Umberto Nencha, Centre de la Mémoire, Service de Gériatrie, Département de Gériatrie et Réadaptation, Hôpitaux Universitaires de Genève, rue Gabrielle Perret-Gentil 6, 1205 Genève, Switzerland. U. Nencha and I. Rigoni share equal contributions.

## Abstract

We assessed the relationship of gamma oscillations with tau deposition in Alzheimer’s disease (AD) and other cognitive diseases, as both are altered during the disease course and relate to neurodegeneration. We retrospectively analyzed data from 7 AD, tau positive patients and 9 tau negative patients, that underwent amyloid PET, and cerebral tau PET and EEG within 12 months. Relative gamma power was higher in tau positive (AD) patients than in tau negative patients (p<.05) and tau burden was associated with a linear increase in gamma power (p<.001), while no association was present with amyloid-β burden. Thus, increase in the gamma power might represent a novel biomarker for tau driven neurodegeneration.

## 1. Introduction

Alzheimer’s Disease (AD) is the most prevalent cause of dementia worldwide (Anon 2021) and its prevalence is expected to increase during the next years. The hallmarks of AD are progressive amyloid-β (Aβ) plaques and neurofibrillary tangles (tau) deposition, and the latter correlates with clinical symptoms, disease severity and speed of progression (Digma et al. 2019; Pontecorvo et al. 2017). Currently, the only way to assess in-vivo tau deposition is through either lumbar puncture or tau PET. In AD, gamma oscillations (>30Hz) show early alterations in the disease course (Verret et al. 2013; Wang et al. 2017). Here, we explored the alterations in the gamma band as proxies of cortical tau deposition in a memory clinic population.

## 2. Methods

### 2.1 Patients

We analyzed data of 7 AD patients with significant amyloid and tau deposition and 9 patients with either a subjective cognitive decline or objective cognitive deficits without tau deposition from the Geneva Memory Center. We included patients who underwent cerebral tau PET and a low-density resting-state EEG within 12 months. This study was approved by the local ethic committee and has been conducted in accordance with the principles of the Declaration of Helsinki (PB_2016-01382 (15-206)).

### 2.3 EEG acquisition and analysis

We selected 5 minutes of the clinical low-density resting state EEG (eyes-opened, 25-channel, sampling rate = 256 Hz). The signal was band-pass filtered ([1-100 Hz]) and notch-filtered at 50 and 100 Hz. Sensors with poor signal-to-noise ratio were removed and interpolated. Independent component analysis and further visual inspection was performed to remove eye blinks and muscle artifacts, after which data were re-referenced to the average. The remaining signal was cropped into epochs of 1 second without overlap (Table 1). The power spectral density was calculated using a single Hanning taper and, for each EEG channel, the relative power in gamma band was defined as the percentage of the power between 30-98 Hz with respect to the total power. For statistical analyses, we used both the relative gamma power calculated for each channel and also averaged over the 25 sensors. EEG analyses were carried out in Matlab.

**Table 1:** Patients’ characteristics and summary. Summary of patient demographic and clinical data. Significant values are highlighted in bold. Summary results are reported as median values [inter-quartile range].

|  | **Tau negative (N=9)** | **Tau positive (N=7)** | **p-value** |
| --- | --- | --- | --- |
| **Age, years** | 73 [70-76] | 71 [69-72.5] | 0.339 |
| **Gender (female), n (%)** | 1(11%) | 7(100%) | **0.002** |
| **Education, years** | 14 [11-16] | 12 [11.5-16.5] | 0.957 |
| **Clinical dementia rating scale** | 0.5 [0-1] | 1 [0.75-1] | 0.094 |
| **Mini mental state examination** | 27 [26-29] | 23 [20-24] | **0.014** |
| **Amyloid positivity, n (%)** | 1 (11%) | 7 (100%) | **0.002** |
| **2-sec EEG epochs retained** | 281 [260 - 292] | 265 [230 - 295] | 0.758 |

### 2.4 PET images acquisition and analysis

PET images were realized using a Siemens Biograph mCT or Vision PET scanner. Cerebral tau-PET images were acquired using 18F-Flortaucipir and amyloid-PET images were acquired using ^18^F-Flutemetamol (N=14) or ^18^F-Florbetapir (N=2). Global tau-PET SUVr was computed as previously described (Mishra et al. 2017), while tau and amyloid status (positive or negative) were assessed visually by an expert nuclear medicine physician (GV) following validated recommendations (Fleisher et al. 2020).

### 2.6 Statistical Analyses

Demographical and clinical data across tau-positive (tau-P) and tau-negative (tau-N) patients were assessed using Mann-Whitney U and Chi-squared test for continuous and categorical variables, respectively. The relative gamma power (averaged over electrodes) was compared with a two-sided Mann-Whitney U-test, and the result was used to define the directionality of a one-sided cluster-based permutation test (Fieldtrip, 2000 permutations). The latter was used to identify clusters of electrodes that, across tau-P patients, showed a greater level of relative gamma power than the tau-N ones. We excluded P9 and P10 as their projections are not described in Okamoto et al. and ran local correlations for the remaining 13 electrodes. For Fz, Cz and Pz, we used the average gamma power of each preponderant cortical area for each hemisphere (e.g. F3/F4 for Fz). We repeated these analyses also for the delta [1-3 Hz], theta [4-7 Hz], alpha [8-12 Hz] and beta [13-29 Hz] bands. Values of relative gamma power were then used for correlation analyses, using a Spearman or a Pearson Correlation Coefficient. For each group (tau-P: N=7; tau-N: N=9) we did the following analyses. 1) We investigated the relationship of average relative gamma power with the global tau-PET SUVr and the amyloid-PET centiloid. 2) For electrodes belonging to the significant cluster, we correlated the local relative gamma power with the SUVr of the region of interest underlying the electrode placement, according to published guidelines (Okamoto et al. 2004). The reported p-values are Bonferroni corrected for the number of electrodes tested.

## 3. Results

Of the 16 selected patients, 7 had a significant cortical tau deposition (i.e. tau-P), while 9 did not (i.e. tau-N). All tau-P patients also showed amyloid deposition on cerebral amyloid-PET, confirming the presence of AD pathology. Among tau-N patients, only one showed amyloid deposition on amyloid-PET. Demographic data and patients’ characteristics are summarized in Table 1.

### 3.1 Gamma oscillations are increased in presence of cortical tau deposition, but not in the presence of amyloid

The average relative gamma power was higher in tau-P than in tau-N patients (p=.042, Fig 1A). Correlation analyses revealed a strong positive correlation between the average gamma power and tau PET SUVr in tau-P patients (rho_Pearson_=0.88, p=0.018, Fig 1B), but not in tau-N (rho_Spearman_=0.7, p=0.087, not shown). No significant correlation was found between the average relative gamma power and the global amyloid burden in neither group (not shown).

**Figure 1:**
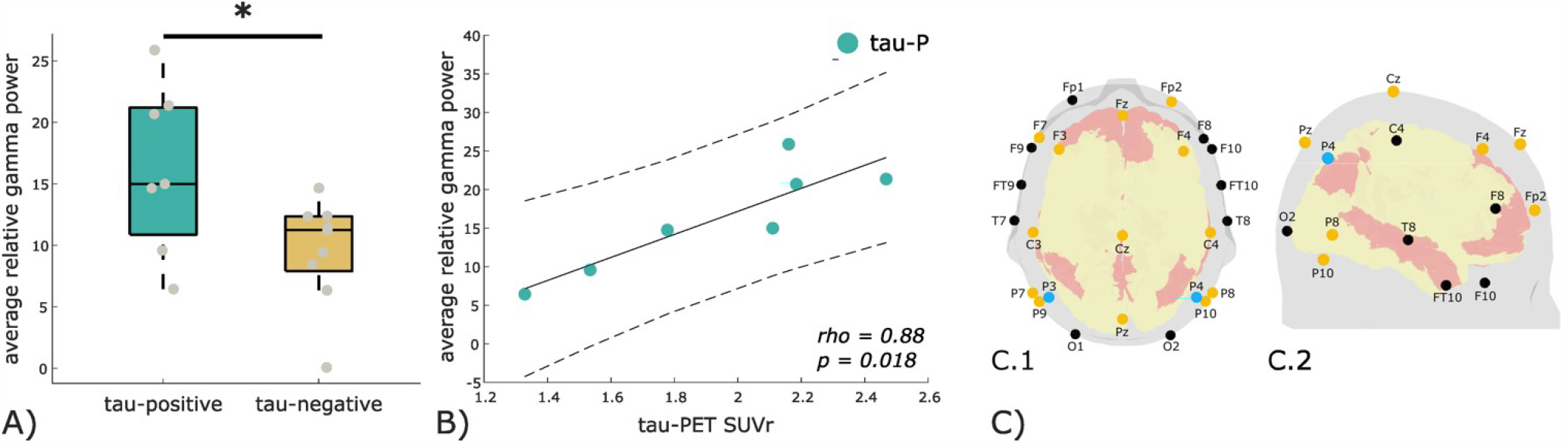
Gamma power in tau-positive and tau-negative patients. A) Boxplot of the average relative gamma power (30-98 Hz) in tau-positive and tau-negative patients. The asterisk indicates statistical significance (p<.05). B) Scatterplot of Pearson correlation between the global tau-PET SUVr and the average gamma power in tau-P patients. C.1) and C.2)) Maps of EEG electrodes (dots) plotted in axial (1) and right lateral sagittal (2) views of the scalp and brain. Yellow dots indicate electrodes that belong to the significant cluster (p<.01). Blue dots indicate the electrodes where local gamma power linearly correlated with local SUVr. Pink areas in are cortical regions belonging to the default mode network (DMN) as previously described (Raichle 2015) adapted to the scale 2 Lausanne parcellation scheme (Cammoun et al. 2012; Hagmann et al. 2008).

### 3.2 Gamma oscillations are increased in frontal and parietal electrodes

Tau-P showed higher gamma power in than in tau-N patients in frontal and parietal electrodes (Fp2, Fz, F3, F4, F7, Cz, C3, C4, Pz, P3, P4, P7, P8, P9, P10, p=.009, Fig 1C). Moreover, in tau-P patients, a positive linear correlation between relative gamma power and local SUVr was found at the parietal electrodes (rho_Pearson_=0.93, p=0.03 and for P3 and rho_Pearson_=0.95, p=0.016 for P4 respectively). No difference is found for the other frequency bands between the tau-positive and tau-negative patients.

## 4. Discussion

EEG alterations have been observed in a large cohort of preclinical AD patients (Gaubert et al. 2019), advancing the hypothesis of complex compensatory mechanisms. Here, we found that gamma oscillations are increased in tau positive patients. We also find a positive linear correlation between relative gamma power and tau (rho=.79, p=.0005), while no correlation was found in other frequency bands nor for amyloid, suggesting a certain specificity of gamma oscillations with tau-related neurodegeneration. Interestingly, gamma oscillations also locally correlate with tau in parietal electrodes. These overlie the parietal and the precuneus cortices, which are known to show early tau-driven neurodegeneration in the course of AD (Frontzkowski et al. 2022). Tau protein accumulation is known to promote network hyperexcitability and in an AD murine model its reduction resulted in a decreased hyperexcitability, independently from the presence of Aβ42 (Tok, Ahnaou, and Drinkenburg 2022). Moreover, a recent study in AD patients found a positive linear correlation between the amplitude of the TMS-evoked potential (TEP) at the level of the posterior parietal cortex and the levels of tau and p-tau in CSF(Casula et al. 2022). Hence, we hypothesize that the presence of tau protein and neurodegeneration could locally disrupt of the physiological rhythms, forcing the remaining neurons to increase their activity, to maintain the excitatory/inhibitory balance. Among the main limitations of this study, except the small sample size due to its retrospective and exploratory nature, we acknowledge the imbalance in sex between tau-P and tau-N, which might have biased some of the analyses, even though to our knowledge gamma oscillations are not known to be influenced by sex.

The lower cognitive scores of tau-P patients on Mini Mental State Examination are in line with increased rate of neurodegeneration expected for these patients. In this view, further longitudinal studies in larger, balanced and age-matched cohorts employing high-density EEG will be crucial to determine the value of gamma oscillations to assess cortical tau deposition.

## Data Availability

Data included in the manuscript are not sharable as patients did not consent to it.

## Abbreviations

AD: Alzheimer’s disease
PET: positron emission tomography
SUVr: standard uptake value ratio
EEG: electroencephalography
Aβ42: amyloid-beta 42

